# A Simple Early Warning Signal for COVID-19

**DOI:** 10.1101/2020.04.28.20083261

**Authors:** Lidia Ceriani, Carlos Hernandez-Suarez, Paolo Verme

**Author notes:** Corresponding Author. This work is part of the program “Building the Evidence on Protracted Forced Displacement: A Multi-Stakeholder Partnership”. The program is funded by UK aid from the United Kingdom’s Department for International Development (DFID), it is managed by the World Bank Group (WBG) and was established in partnership with the United Nations High Commissioner for Refugees (UNHCR). The scope of the program is to expand the global knowledge on forced displacement by funding quality research and disseminating results for the use of practitioners and policy makers. This work does not necessarily reflect the views of DFID, the WBG or UNHCR.

## Abstract

The paper provides some initial evidence that daily mortality rates (for any cause) by municipality or province can be used as a statistically reliable predictor of looming COVID-19 crises. Using recently published deaths figures for 1,689 Italian municipalities, we estimate the growth in daily mortality rates between the period 2015–2019 and 2020 by province. All provinces that experienced a major COVID-19 shock in mid-March 2020 had increases in mortality rates of 100% or above already in early February 2020. This increase was particularly strong for males and older people, two recognizable features of COVID-19. Using a panel fixed effect model, we show that the association between these early increases in mortality for any cause and the March 2020 COVID-19 shock is strong and significant. We conclude that the growth in mortality rates can be used as a statistically reliable predictor of COVID-19 crises.

## 1 Introduction

It is now almost certain that the spread of COVID-19 and the growth of COVID-19 related deaths emerged much earlier than previously thought [3]. Recent studies have also argued that the lethality of the virus among the population at large is not high, in fact similar to the seasonal flu [7]. However, in some geographical areas and for some population groups, the virus has been extremely morbid and lethal to an extent that emergencies rooms, hospital admissions and beds, intensive care units and even funeral homes have been overwhelmed and incapable of responding to demand. This phenomenon also occurs over a very short period of time. From the time authorities first observe a rapid growth in infections and deaths to the time the health system collapses only one to three weeks pass, a very short window of time to prepare local institutions for a proper response. This, in turn, contributes to the failure of the health system and the growth in deaths.

A very different outcome could be achieved if local and national authorities were able to predict the peak of the crisis three-four weeks in advance. This would provide significantly more time to prepare emergency rooms, hospital admissions and beds, intensive care units and stock up with essential medical supplies. This paper argues that this is possible by simply monitoring changes in daily mortality rates in real time. We illustrate this claim with newly released data on deaths by municipality in Italy. A recent article on the Financial Times shows that these data are available for many countries [2].

The paper shows that (i) the mortality rate in Italy (for any cause) was rising sharply and deviating significantly from previous years’ average in many Italian provinces already in January and February 2020 and (ii) growth in these mortality rates by province can be used as a statistically reliable early warning indicator to identify those provinces heading towards a collapse of the health system due to COVID-19. This indicator alone can anticipate a COVID-19 crisis by several weeks. Had this information been available to policy makers in mid-February 2020 and had the lock-down in Italy been introduced at the same time, many lives could have been saved. As COVID-19 is still spreading across the world, we recommend to monitor the growth in mortality rates in real time and use a doubling of the mortality rate as a rule of thumb to reliably predict the administrative areas that are heading towards a COVID-19 crisis.

The next section illustrates the data, section 3 provides the analysis of the mortality rates by province, section 4 shows how the mortality rate can be used as a predictor for municipalities heading towards a crisis and section 5 discusses results.

## 2 Data

The number of daily deaths is a mandatory indicator that all municipalities in Italy are required to provide to various central administrations, including the Italian National Institute of Statistics *(Istituto Italiano di Statistica* – ISTAT). This information is validated, elaborated and provided publicly by means of *i.Istat*, the official national repository of statistical data maintained by ISTAT. This process is lengthy and, under normal circumstances, ISTAT provides this information publicly one to two years following reception of these data. Under the exceptional circumstances of COVID-19, ISTAT has been requested by the Ministry of Health to produce and publish these data rapidly and has recently released the first figures for the months of January-April 2020 for a selection of municipalities [4]. We analyze these data to understand whether they could have been used to anticipate the health crisis experienced by many provinces in Italy during the month of March 2020.

We define a “crisis” as a two-times’ fold growth in mortality rates (200% growth rate) or above as compared to previous years. As we will show, the Italian provinces that experienced a major COVID-19 shock did so around the period March 10th-20th. During this period and in these provinces, the average mortality rate was over four folds higher than in previous years. This explains why local administrations could not cope in terms of response, from hospital beds to funeral services.

The data provided by ISTAT include the daily number of deaths for 1,689 municipalities. These municipalities are not a representative sample of Italian municipalities and have been selected by ISTAT following three criteria: (i) the completeness and timely recollection of information; (ii) number of deaths increasing more than 20 percent in the period March 1st–April 4th 2020 in comparison to the average over the same period for the years 2015–2019; (iii) number of deaths in the period January 4th-April 4th 2020 of at least 10. Data are also provided by gender and age group (22 age groups, in brackets of 5 years each, with the exception of the two groups made of individuals age < 1 and > 100). These data focus therefore on the municipalities that have experienced the largest increases in the number of deaths and allow for an analysis by gender and age, two key dimensions to understand whether the growth in deaths for any cause relates to COVID-19 deaths.

From these data, we first computed mortality rates as the number of deaths over the the population using population figures available at i.ISTAT [5]. For a given location *l*, gender *g*, and age group *a*, the mortality rate at time *t* is defined as:

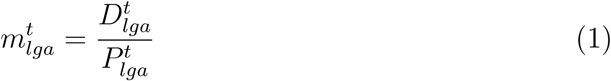

where 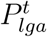 is the population size, 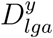 indicates the number of deaths and *t* is time expressed in days. The growth rate in the mortality rate between the period 2015 and 2019 and 2020 is then defined as:

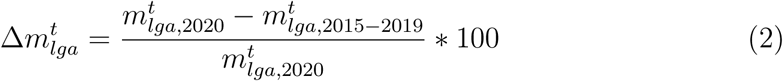

To compute these rates, we take the resident population by gender and age as of January 1st of each year for the years 2015–2019. For 2020, we impute the same population size as of 2019, under the hypothesis that there is zero population growth in 2020. This should not affect results as the population of Italy has been very stable during the past five years and is not expected to increase significantly in 2020.

We further cleaned the dataset to remove all instances where a municipality did not report any data, and all observations pertaining to two new municipalities instituted in February 2019 for which population data were not available (Lu e Cuccaro Monferrato, and Valbrenta). We also dropped observations on the number of deaths reported for February 29th, as only two of the six years under consideration are leap years (2016 and 2020).

Although data are available at the municipality-level, the analysis is run at the level of provinces. These are the intermediate administrative divisions between municipalities and regions. We do so to smooth municipalities figures (daily figures on mortality at the municipality level contain many zeroes), and because the coverage areas of hospitals is more likely to be at the provincial level. Hospitals that have been overwhelmed by demand in mid-March 2020 cover areas that are larger than the municipality they reside in. They typically cover provinces and, in some cases, they cover regions.^1^ The analysis is performed using STATA. To avoid major daily fluctuations, data were also smoothen using the STATA function `lowess` [9], which transforms data using a locally weighted regression (bandwidth of 0.8). Following data cleaning and smoothing, the final data set included 9,419 observations. This is a panel of daily observations for the period January 1st and April 4th, 2020 covering 103 provinces. The panel is not balanced as some provinces miss some days.

## 3 Growth in Mortality

A growth in daily mortality rates (any cause) was already very visible for many Italian provinces in early February 2020.^2^ Figure 1 shows the growth rate in the mortality rates as defined in Equation (2) between January 1st and April 4th, 2020 for five of the provinces that became known in March 2020 for their COVID-19 mortality crisis (Bergamo, Cremona, Lodi, Piacenza and Brescia). A percent increase of 100 on the *y-axis* indicates that the mortality rate has doubled in 2020 as compared with the average of the period 2015–2019. It can be clearly seen that the growth rate of mortality starts to deviate from zero already in early February and doubles around mid-February 2020.

**Figure 1:**
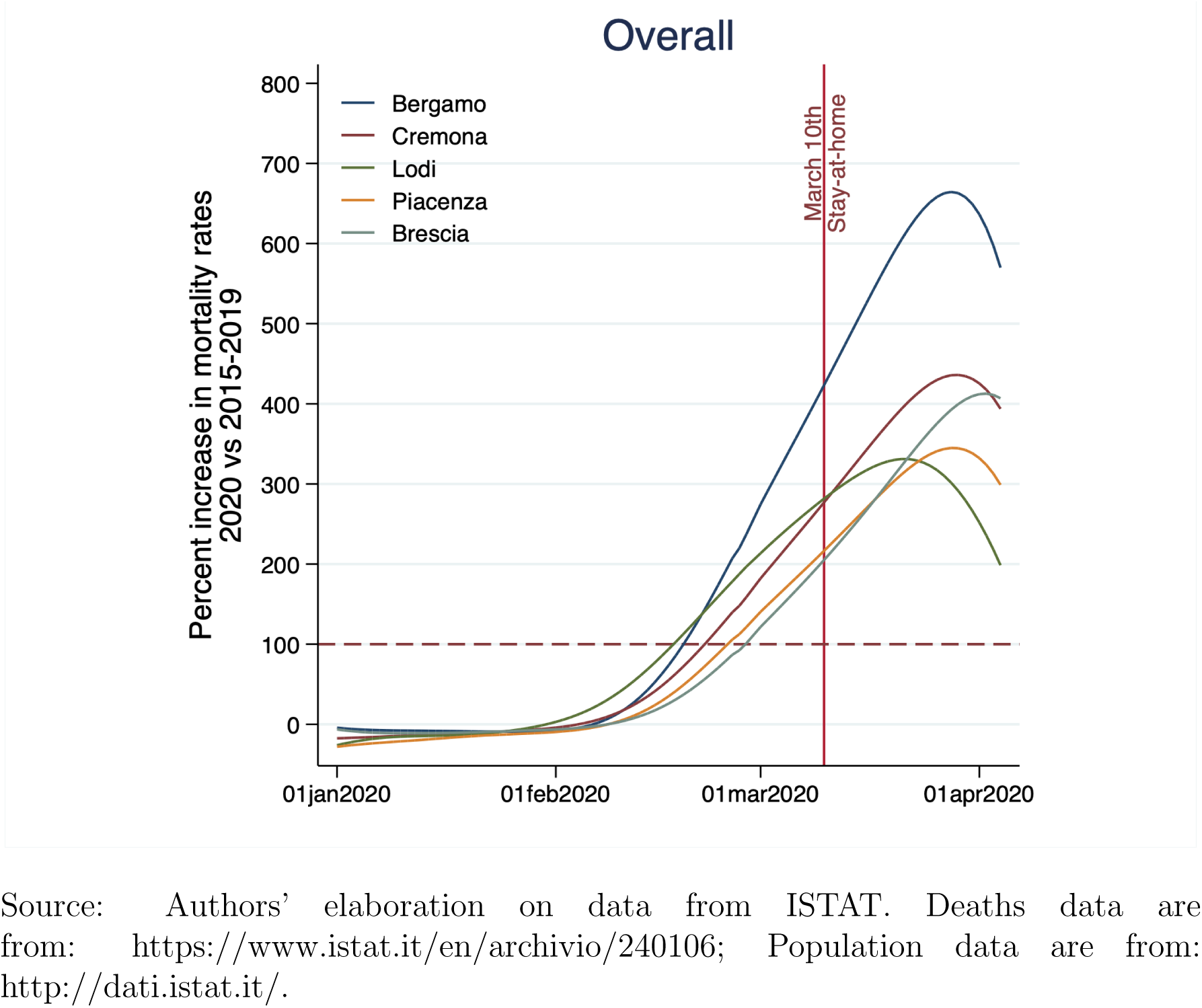
Growth in Mortality Rate 2020/2015–2019

These same figures show that the growth in mortality rate (any cause) was particularly sharp for men and old people. Figure 2 shows the same curves shown in Figure 1 by gender. The figure shows that the curves for males are steeper and have increased to higher levels than those for females by March 2020, especially in the hardest hit provinces such as Bergamo. Figure 3 repeats the exercise by age groups and shows that the mortality crisis has been remarkably sharp for people age 60 and above, in fact this age group exhibits a growth in mortality that is more than twice as high as the one for the age group below the age of 60.

**Figure 2:**
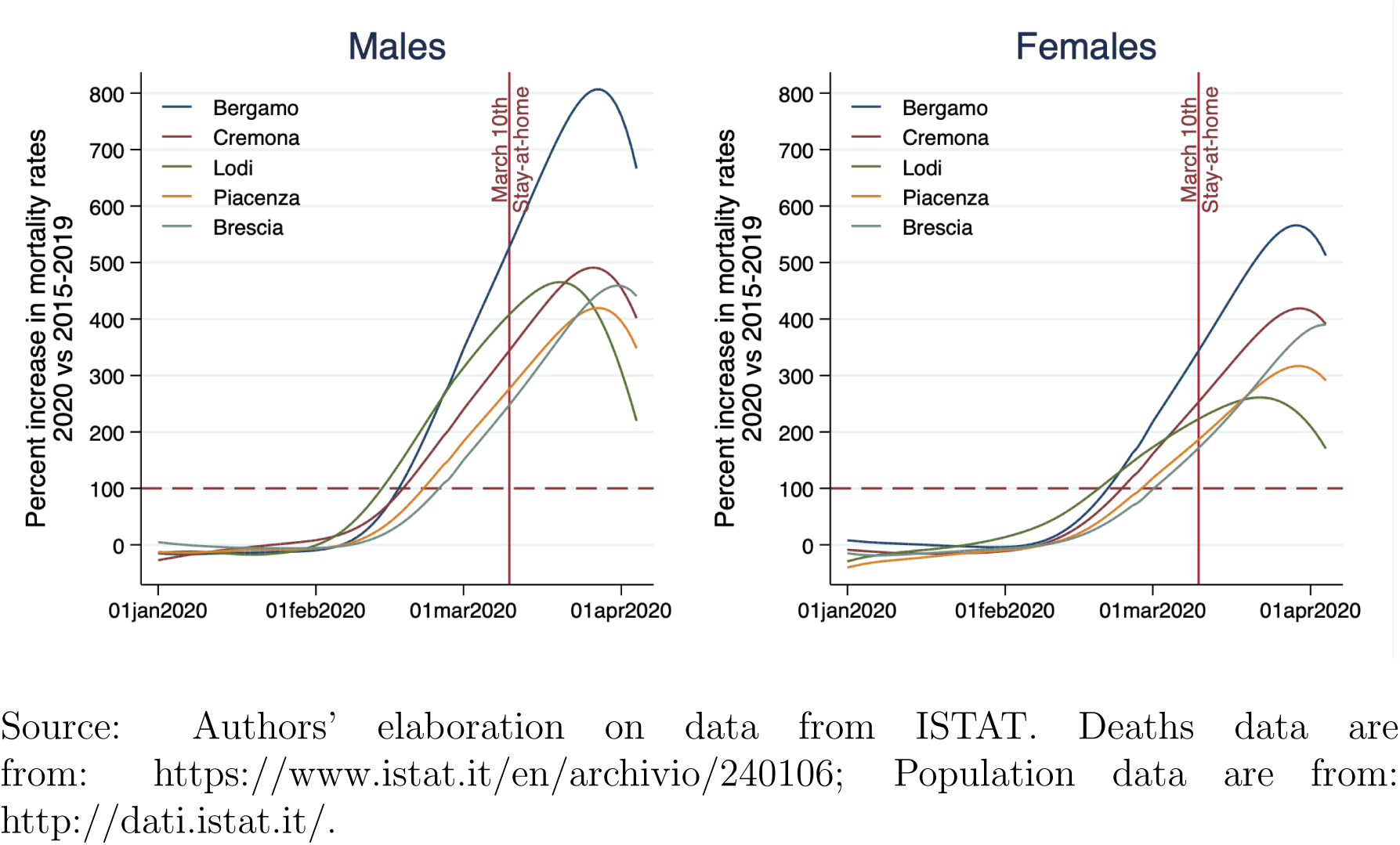
Growth in Mortality Rate 2020/2015–2019 – Gender

**Figure 3:**
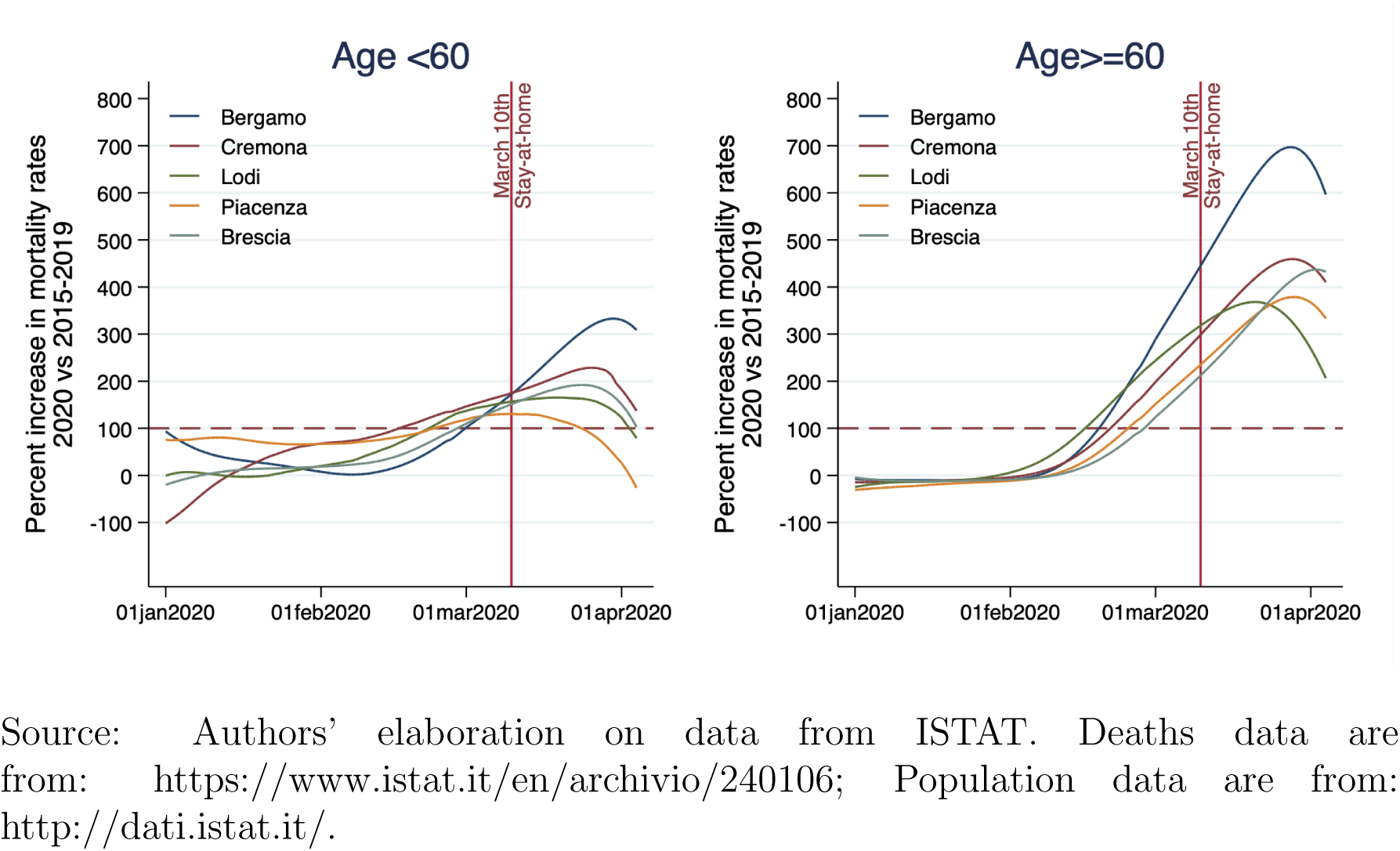
Growth in Mortality Rate 2020/2015–2019 - Age Groups

By splitting the growth in mortality rates by gender and age, it is possible therefore to quickly recognize the deadly characteristics of COVID-19, even in the absence of data on the causes of deaths. This is important because the data on the causes of deaths are complex as many deaths are cataloged with multiple causes while others are mis-classified, which was probably the case for the early deaths due to COVID-19. In essence, plotting these simple graphs by province in mid-February 2020 could have provided an indication where the health crisis was going to unfold. But could these figures be used to make statistically reliable predictions? The next section addresses this question.

## 4 Predicting the crisis

We show with a simple panel equation how the sharp growth in mortality experienced by many Italian provinces during the period March 10th-20th, 2020 could have been predicted by simply observing the growth in mortality rates during the months of January and February, 2020.

Recall that we dispose of a panel data set of mortality rates growth for Italian provinces with daily observations on the number of deaths covering the period January 1st - April 4th for each year between 2015 and 2020. With these data, we can estimate a panel prediction model where we use as predictors lagged variables of the growth in mortality rate. The model is described as follows:

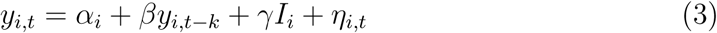

where *i* and *t* are provinces and time expressed in days respectively, *y_i_,_t_* is the growth of the mortality rate in 2020 as compared to the period 2015–2019, *y_i_,_t−k_* is this same variable lagged by *k* days, *I_i_* are provinces fixed effects and *η_it_* is the error term. This is what is generally referred to as a panel fixed effects model. Similar models which are popular for this kind of analysis are the Arellano-Bond types of models [1].

In our case, the outputs of interest are both *β* and *γ*. The *β* coefficient will provide evidence on how long we can go back in time to find growth in mortality rates that are correlated with the present growth in mortality. The *γ* coefficients will tell us instead which provinces are more likely to have higher mortality growth with such lags. And the significance level of the *γ* coefficients informs us on how strong the prediction of higher growth in mortality is. We also compare these coefficients with the average mortality growth rate for the period March 10th-20th, which is the period where mortality peaked. With this comparison, we are able to tell for how many provinces we could have predicted in early February 2020 the sharp growth in mortality observed in mid-March 2020.

Table 1 shows the results for the top ten provinces that experienced a growth rate in mortality of over 200% in March 10th-20th. Results are sorted using the average mortality rate growth for the same period to see whether the model also ranks provinces in order of magnitude of the mortality shock. We find significant fixed effects for all but one of the top ten provinces that experienced a mortality growth rate of over 200% in March 10th-20th (90% accuracy), with this result applying to all three time lags considered (7, 14 and 21 days). Also, the ranking of provinces according to the significance level (*z − stat*) is very similar to the rank in terms of depth of crisis in mid-March 2020. In fact, all provinces are ranked correctly with only two exceptions.

**Table 1:** Panel Fixed Effects Equations and the Average Mortality Rate Growth (MRG) During the Crisis

| Province | Lag 7 |  | Lag 14 |  | Lag 21 |  | MRG |
| --- | --- | --- | --- | --- | --- | --- | --- |
|  | Ceff. | z-stat. | Ceff. | z-stat. | Ceff. | z-stat. | March 10-20 % |
| MRG Lagged | 0.4 | 37.2 | 0.3 | 20.9 | 0.1 | 6.7 |  |
| Lombardia_Bergamo | 138.5 | 10.2 | 185.7 | 12.4 | 231.5 | 14.4 | 755.4 |
| Lombardia_Cremona | 93.5 | 7.0 | 123.6 | 8.3 | 154.8 | 9.6 | 475.7 |
| Lombardia_Lodi | 87.0 | 6.5 | 116.9 | 7.8 | 148.1 | 9.2 | 445.7 |
| Emilia-Romagna_Piacenza | 71.2 | 5.3 | 94.7 | 6.4 | 116.2 | 7.2 | 400.2 |
| Lombardia_Brescia | 70.9 | 5.3 | 95.0 | 6.4 | 114.0 | 7.1 | 320.4 |
| Abruzzo_Pescara | 30.5 | 2.3 | 39.4 | 2.6 | 57.2 | 3.5 | 299.2 |
| Veneto_Belluno | 22.7 | 1.7 | 27.4 | 1.8 | 41.4 | 2.6 | 297.7 |
| Emilia-Romagna_Parma | 51.1 | 3.8 | 65.0 | 4.4 | 80.5 | 5.0 | 284.3 |
| Sicilia_Enna | 37.2 | 2.8 | 47.0 | 3.1 | 54.5 | 3.4 | 255.6 |

Source: Authors' elaboration on data from ISTAT. Deaths data are from: https://www.istat.it/en/archivio/240106; Population data are from: http://dati.istat.it/.

## 5 Discussion

Using recently released data of deaths for any cause covering the period January-April, 2020, we showed that the growth in daily mortality rates was already above 100% in early February 2020 in many Italian provinces. We also showed that these provinces are the same that experienced a major spike in mortality due to COVID-19 in mid-March, 2020. Using a simple panel model, we then showed that there is a robust statistical association between the growth in mortality rate and its lagged values of one, two or three weeks. Simply monitoring the daily growth rate in mortality rate by province would have shown that some provinces were experiencing a major shock in early February, 2020, an indication that could have alerted and mobilized the Italian authorities three to four weeks in advance of the lock-down day of March 10th, 2020.

The natural question that arises is why this indicator was not being monitored in January and February 2020. Some local municipalities may have noticed an increase in deaths but such increase could not be properly compared with previous years and other municipalities and was not sufficiently sharp to overwhelm local services such as hospitals or funeral homes in January or February, 2020. Local authorities were not yet observing dysfunctions in public services and may have attributed the increase in deaths to a random spike. Also, in our understanding, the growth in daily mortality rate by municipality or province was not an indicator included in the monitoring system of the flu. Italy has a rather sophisticated monitoring system for the flu (Influnet) which collects information from doctors, laboratories and the population on a weekly basis to monitor the development of flu epidemics in real time. These indicators are reviewed on a weekly basis and constitute the main source of information for a weekly report prepared by the flu surveillance system [6]. In our knowledge, this system does not integrate information on growth in mortality rates for any cause, possibly because it focuses on the flu only.

If Italian authorities had monitored the growth in mortality rates in real time, it would have been immediately obvious that something very atypical was occurring. While the growth in mortality rate may not be a standard monitoring indicator for the seasonal flu, we showed that it can be a very effective indicator to alert authorities of unfolding COVID-19 health crises. We recommend therefore to monitor this indicator in real time and, as a rule of thumb, use a 100% increase in the death rate at the provincial level as an early warning signal. In particular, generating the growth rate in mortality by age and gender can provide some early indications that this growth is related to COVID-19. This is especially important in view of the current COVID-19 crisis, which is expected to re-surge in countries where the lock-down will be eased, or is still in its early phase as in many countries around the world.

## Data Availability

All data used in the manuscript are publicly available on-line free of charge.

https://www.istat.it/en/archivio/240106

http://dati.istat.it/

## Footnotes

1 The Italian health system is decentralized and the administrative level that is responsible for health services is the region.

2 See also [8] for an illustration of an earlier version of these data.

## Notes

### Competing Interest Statement

The authors have declared no competing interest.

### Funding Statement

The study was funded by UK aid from the United Kingdom's Department for International Development (DFID). A full disclaimer is on the first page of the manuscript.

